# A first-in-human evaluation of the safety and immunogenicity of SCB-2019, an adjuvanted, recombinant SARS-CoV-2 trimeric S-protein subunit vaccine for COVID-19 in healthy adults; a phase 1, randomised, double-blind, placebo-controlled trial

**DOI:** 10.1101/2020.12.03.20243709

**Authors:** Peter Richmond, Lara Hatchuel, Min Dong, Brenda Ma, Branda Hu, Igor Smolenov, Ping Li, Peng Liang, Htay Htay Han, Joshua Liang, Ralf Clemens

**Affiliations:** Division of Paediatrics, University of Western Australia, Wesfarmers Centre of Vaccines and Infectious Diseases, Telethon Kids Institute and Perth Children’s Hospital, Perth, Western Australia; Linear Clinical Research, Nedlands, Western Australia; Clover Biopharmaceuticals, Chengdu, China; Global Research in Infectious Diseases (GRID), Rio de Janeiro, Brazil

## Abstract

**Background:** As part of the accelerated development of prophylactic vaccines against severe acute respiratory syndrome coronavirus 2 (SARS-CoV-2) we report a first-in-human dose-finding and adjuvant justification study of SCB-2019, a novel protein subunit vaccine candidate composed of a stabilised trimeric form of the spike (S)-protein produced in CHO-cells, combined with two different adjuvants.

**Methods:** This phase 1 study was done in one centre in Western Australia in 151 healthy adult volunteers in two age groups (18–54 and 55–75 years), allocated to 15 groups (nine young and six older adults) to receive two doses, 21 days apart, of placebo, or 3 μg, 9 μg or 30 μg SCB-2019, alone or adjuvanted with AS03 or CpG/Alum. Reactogenicity was assessed for 7 days after each vaccination. Humoral responses were measured as SCB-2019 binding and ACE2-competitive binding IgG antibodies by ELISA, and as neutralising antibodies by wild-type SARS-CoV-2 microneutralisation assay; cellular responses to pooled S-protein peptides were measured by flow-cytometric intracellular cytokine staining.

**Findings:** We report on 148 participants with at least 4 weeks follow-up post dose 2. Three participants withdrew, two for personal reasons and one with an unrelated SAE (pituitary adenoma). Vaccination was well tolerated, with few Grade 3 solicited adverse events (AE). Most local AEs were mild injection site pain, which were more frequent with formulations containing AS03 than CpG/Alum or unadjuvanted SCB-2019. Systemic AEs, mostly transient headache, fatigue or myalgia, were more frequent in young adults than older adults after the first dose, but similar after second doses. Unadjuvanted SCB-2019 elicited minimal immune responses, but SCB-2019 with fixed doses of AS03 or CpG/Alum induced high titres and seroconversion rates of binding and neutralising antibodies in both young and older adults. Titres were higher than those observed in a panel of COVID-19 convalescent sera in all AS03 groups and high dose CpG/Alum groups. Both adjuvanted formulations elicited Th1-biased CD4+ T cell responses.

**Interpretation:** SCB-2019 trimeric protein formulated with AS03 or CpG/Alum adjuvants elicited robust humoral and cellular immune responses against SARS-CoV-2 with high viral neutralising activity. Both adjuvanted formulations were well tolerated and are suitable for further clinical development.

**Clinical trial registration:** ClinicalTrials.gov identifier NCT04405908.

## INTRODUCTION

To date, the global COVID-19 pandemic due to the SARS-CoV-2 virus has caused 65 million infections and almost 1·5 million deaths [1]. Infections are leading to unprecedented numbers of cases of severe respiratory illness with significant proportions of patients requiring admission to intensive care units (ICU) [2]. COVID-19 is associated with a high transmission rate and without adequately effective therapies rising numbers of cases of respiratory distress are threatening to overwhelm global healthcare capacity. Interventions are urgently required to reduce this disease burden leading to the accelerated introduction into clinical development of at least 47 vaccine candidates [3].

The main viral antigenic target is the glycosylated Spike (S) protein, a trimeric protein consisting of two subunits, S1 and S2 [4], which is an essential component for viral binding, fusion and uptake into mammalian cells [5]. S1 interacts with the receptor binding domain (RBD) of human cell-surface human angiotensin-converting enzyme 2 (ACE2) and following proteolytic cleavage of the two subunits the S2 domain undergoes a major conformational change which leads to fusion and intracellular uptake of the viral mRNA for replication [6,7]. Interference with this process is the basis of most immunological approaches to prevent COVID-19 infections including vaccines [8].

Trimer-Tag^©^ is derived from the C-terminal region of human type I procollagen and is capable self-trimerization [9]. When soluble receptors or biologically active proteins are fused in-frame to Trimer-Tag^©^, the resulting fusion proteins expressed in mammalian cells are secreted as disulphide bond-linked homotrimers. SCB-2019, a recombinant SARS-CoV-2 S-Trimer fusion protein produced in Chinese hamster ovary (CHO) cells, preserves the native trimeric structure of S-protein in the prefusion form. SCB-2019 binds with high affinity to human ACE2 [10]. When formulated with the oil-in-water emulsion adjuvant, AS03, or the TLR9 agonist, CpG, combined with Alum [10], SCB-2019 induced protective immunity to SARS-CoV-2 *in vivo* in animal challenge studies [11]. This first-in-human phase 1 dose-finding and adjuvant justification study was performed to assess the safety, tolerability and immunogenicity of three dose levels of SCB-2019 when administered to healthy adults as two doses 21 days apart without adjuvant or formulated with either AS03 or CpG/Alum.

## METHODS

We report the interim analysis of the first stage of a phase 1 randomized, double-blind, placebo-controlled study of SCB-2019. The trial was done in one study centre in Nedlands, Western Australia, Australia from June 19, 2020 until database lock on October 20, 2020. The study protocol was approved by the study centre Institutional Review Board and registered with ClinicalTrials.gov (identifier NCT04405908). The study was done according to International Conference of Harmonisation and Good Clinical Practice guidelines.

The overall objectives were to assess the safety, tolerability and immunogenicity of three increasing dosages of SCB-2019, unadjuvanted or adjuvanted with AS03 or CpG/Alum, in young and older adults when administered as two intramuscular doses 21 days apart.

### Study design

This first stage was a placebo-controlled dosage escalation study done in two parts with 15 groups of 10 participants each. The first part was done in nine groups of young adults (18–54 years inclusive) using a sentinel strategy in which the first two participants assigned to each group (one vaccinee, one placebo control) received their study injections. Sentinels were monitored for 48 hours and the safety data reviewed by a Safety Monitoring Committee (SMC) to assess any significant AEs that occurred before the remaining eight participants of that group (seven vaccinees, one placebo) were treated. No sentinel strategy was applied to the second part of the study performed in six groups of older adults (55–75 years inclusive) for which recruitment was only started after the safety from the equivalent adult group (same dose and formulation) had been considered by the SMC. Further expansion of the study is planned for long term safety follow-up as well to generate data on antibody persistence, and to include SARS-CoV-2 seropositive participants. Results of those investigations will be reported separately.

### Participants

Eligible participants were adults of either sex from 18 to 75 years of age who were healthy at enrolment based on medical history and medical assessment. All volunteers were screened for serum antibodies against SARS-CoV-2 as evidence of prior infection, and for acute exposure using reverse transcriptase-polymer chain reaction (RT-PCR), which were repeated at each study visit. Inclusion criteria included being able to provide informed consent, having a BMI between 18·5 and 35·0 kg/m^2^, being able to understand and sign the informed consent and being available for the duration of the study (6 months). Female participants of childbearing potential were not to be pregnant or breastfeeding and had to agree to use protocol-approved forms of contraception until 6 months after the first vaccination. Men were also to use a protocol-approved form of contraception from the day of first vaccination until 6 months after the first vaccination and refrain from donating sperm over the same period.

Main exclusion criteria included positive serology for SARS-CoV-2, any uncontrolled chronic medical disorders, any known or suspected impairment of the immune system due to known immunosuppressive conditions or any therapy with immunosuppressants or immunostimulants, known allergy to any vaccine components, malignancies, a positive screening serology for HIV, hepatitis B or C, or prior receipt of any other SARS-CoV-2 vaccine. All volunteers were asked to avoid strenuous exercise from screening to Day 50.

### Vaccine

SCB-2019 is supplied in single-use vials as a sterile, clear to slightly opalescent and colourless solution for injection. Before use vials were stored at 2°C to 8°C. For the three selected dose levels the appropriate dose of SCB-2019 was diluted in a vial with sodium chloride (0·9%). For adjuvant mixing and administration 0·25 mL of AS03 (GSK Vaccines, Wavre, Belgium) or 1·5mg CpG 1018 (Dynavax Technologies, Emeryville, CA, USA) plus 0·75 mg Alum (Alhydrogel, Croda, Goole, UK) per dose were added to the vial and mixed by gentle inversion at room temperature for administration within 1 hour. Each 0·5 mL vaccine dose, containing 3 μg, 9 μg or 30 μg SCB-2019 with sodium phosphate buffer and 0·05 mg polysorbate 80 in 0·9% sodium chloride, was withdrawn into a syringe for injection and administered by intramuscular injection in the deltoid region. Placebo was 0.5mL 0·9% sodium chloride for injection.

Participants were assigned a study number at enrolment and vaccinated according to a randomisation list prepared by the study sponsor. All participants and personnel involved in safety data collection and immunogenicity assessments were blinded to the study treatment. Vaccine preparation and administration were performed by different unblinded study personnel, using opacified syringes to maintain the participant blind as the vaccine and placebo are visually different.

### Safety assessments

On Day 1, before vaccination, each participant received a full physical examination when vital signs were recorded and a blood sample was drawn for baseline safety laboratory parameters. Further safety blood samples were drawn on Days 8, 22, 36 and 50. Safety laboratory tests included haematology, coagulation panel, serum chemistry, and urinalysis. Following vaccination sentinel subjects were monitored in the study centre for 6 hours, all other participants remained under observation for 60 minutes for potential immediate post-vaccination reactions. Participants then completed daily electronic diary cards to record solicited local reactions (pain, redness and swelling at the injection site), systemic adverse events (headache, fatigue, myalgia, nausea/vomiting, diarrhoea and vomiting) and body temperature for 7 days. Solicited reactions and AEs were graded for severity (*see Supplementary Appendix pages 2–3*) by the participants and assessed for causality by the investigator during interview at the following study visit. All unsolicited AEs were recorded from Day 1 to Day 50. Serious adverse events (SAE) and adverse events of special interest (AESI) (*see Supplementary Appendix page 2*) occurring before the database lock were to be reported immediately to the investigator and then to the study sponsor within 24 hours. During study conduct a safety monitoring committee continuously assessed safety data with the option to authorise use of stopping/pausing rules predefined in the protocol.

All study participants were tested for SARS-CoV-2 by nasopharyngeal swab for reverse transcription polymerase chain reaction (RT-PCR) at each study visit. If a participant was suspected to be infected with SARS-CoV-2 virus or had confirmed COVID-19 between study visits, the participant was requested to have an additional test for SARS-CoV-2 infection.

### Immunogenicity assessments

Blood was drawn to prepare serum samples for immunogenicity assessments before the vaccinations on Days 1 and 22, and then on Days 36 and 50. Sera were stored at ≤-80°C until shipment to the immunological laboratory (360biolabs, Melbourne, Victoria, Australia) for analysis. Four immunological assays were performed to evaluate humoral immune response following the study vaccination. The primary immunogenicity endpoint was based on the anti-SCB-2019 IgG antibody titre at each blood sampling timepoint, measured by ELISA. Secondary immunogenicity assessments included an ACE2-competitive ELISA measuring the inhibition of SCB-2019 binding to human ACE2 receptor by serum antibodies, and anti-wild-type SARS-CoV-2 neutralising activity measured by wild-type microneutralisation assay (WT-MN_50_). A panel of 20 human CPVID-19 convalescent serum samples from three hospitalised and 17 non-hospitalised adults (mean age 37 years; sera collected 20-57 days after symptom onset; mean 39 days) and a NIBSC reference serum 20/130 were analysed using the same validated assays as comparators for the post-vaccination serum samples. Details of the immunological assay methods are provided in Supplementary Materials (*Supplementary Appendix pages 4–5*).

PBMCs were collected from all participants at Days 1, 22, 36 and 50 to assess T-cell mediated immune responses to vaccination using intracellular cytokine staining (ICS) flow cytometry to measure CD4-positive T cells expressing markers including IFN-γ, IL-2, IL 4, IL-5 and IL-17 after stimulation with SARS-CoV-2 S-protein peptide pools.

### Statistics

There was no formal statistical hypothesis in this phase 1 study and all data summaries are presented descriptively by group. The study sample size was not based on any statistical hypothesis but is typical of such phase 1 studies and was considered to be adequate to provide a preliminary assessment of vaccine safety and reactogenicity in each cohort.

The Safety Analysis Set (SAS) consists of all subjects randomised to receive at least one dose of study vaccine or placebo, analysed according to the treatment they actually received. Reported summary statistics include counts and percentage of participants who reported at least one solicited local reactions and systemic AEs and unsolicited AEs (with severity and causality), and SAEs and AESIs, after the first and second doses. For this report, the safety data for all participants with at least 21-day safety follow up after Dose 1 are included.

The main analysis population for the immunogenicity analysis in this report is the immunogenicity full analysis set, consisting of all participants in the SAS with at least one post vaccination blood sample collected and analysed for immunogenicity. Subjects are summarised according to treatment received. Antibody responses are presented as geometric mean titres (GMT) with 95% confidence intervals (95% CI) at each blood sampling timepoint for each vaccine group. Geometric mean values are calculated on Log10 (titres/data) values, with subsequent antilog transformations applied, the 95% CI being calculated using normal distribution. Seroconversion rates, defined as the percentage of participants with at least a four-fold increase in antibody titre over baseline within each study group, were calculated for the Day 22, 36 and 50 timepoints. For the between group comparisons in geometric means an analysis of variance (ANOVA) model was fitted to log-transformed assessment (such as titre) values based on the subjects with available data at each timepoint then the geometric mean ratio (GMR) and the 95% CI were calculated. Two-sided 95% CIs for the GMR were obtained by calculating CIs using Student’s t-distribution for the mean difference of the logarithmically transformed results and antilog transformation of the confidence limits. All analyses, and summaries were on group unblinded data performed using SAS^®^ software (version 9·4 or higher) or GraphPad Prism, v.6.0c.

## RESULTS

### Study population

A total of 329 volunteers were screened, of whom 151 (91 young adults and 60 older adults) were enrolled in their respective age strata after testing negative for SARS-CoV-2 (**figure 1**). The majority of the screen failures (173 of 178 [97%]) were due to exclusion criteria. One adult assigned to the 30 μg SCB-2019 group who withdrew before receiving any vaccination was replaced with another volunteer. Demographics were similar across groups (**table 1**). In the young adult groups the mean age of all SCB-2019 recipients was 36·2 ± 11·5 years vs. 32·6 ± 10·7 years for placebo recipients, 40% were male, and most described themselves as white and neither Hispanic nor Latino. In older adults the mean age was 61·1 ± 4·9 years in vaccinees and 62·3 ± 5·9 years in placebo controls, 47% were male, and all were white.

**Table 1.** Demographics of the younger and older adult study populations per vaccine group (placebo groups combined)

|  |  | 3 µg SCB-2019 |  |  | 9 µg SCB-2019 |  |  | 30 µg SCB-2019 |  |  | Placebo |
| --- | --- | --- | --- | --- | --- | --- | --- | --- | --- | --- | --- |
|  |  | Plain | ASO3 | CpG/Alum | Plain | ASO3 | CpG/Alum | Plain | ASO3 | CpG/Alum |  |
| Young adults (18–54 years) |  |  |  |  |  |  |  |  |  |  |  |
|  | N = | 8 | 8 | 8 | 8 | 8 | 8 | 9 | 8 | 8 | 18 |
| Age (yrs) | Mean | 37.6 | 35.8 | 41.9 | 36.5 | 37.0 | 36.1 | 30.9 | 31.3 | 39.1 | 32.6 |
|  | SD | 11.9 | 9.3 | 11.1 | 12.4 | 11.6 | 15.0 | 11.4 | 11.3 | 9.9 | 10.7 |
|  | range | (20–50) | (24–53) | (20–53) | (20–54) | (21–53) | (19–55) | (18–49) | (19–47) | (21–50) | (18–52) |
| Male | n (%) | 5 (63) | 5 (63) | 0 | 4 (50) | 4 (50) | 4 (50) | 3 (33) | 2 (25) | 2 (25) | 7 (39) |
| Race | n (%) |  |  |  |  |  |  |  |  |  |  |
| Asian |  | 2 (25) | 1 (13) | 1 (13) | 1 (13) | 1 (13) | 1 (13) | 1 (11) | 2 (25) | 2 (25) | 4 (22) |
| Black |  | - | - | - | - | - | 1 (13) | - | - | - | - |
| White |  | 6 (75) | 6 (75) | 7 (88) | 6 (75) | 7 (88) | 6 (75) | 8 (89) | 6 (75) | 6 (75) | 14 (78) |
| Other |  | - | 1 (13) | - | 1 (13) | - | - | - | - | - | - |
| Ethnicity | n (%) |  |  |  |  |  |  |  |  |  |  |
| Hispanic or Latino |  | - | - | 2 (25) | 2 (25) | 1 (13) | - | - | 1 (13) | - | 1 (6) |
| Not Hispanic or Latino |  | 8 (100) | 8 (100) | 6 (75) | 6 7(5) | 7 (88) | 8 (100) | 9 (100) | 7 (88) | 8 (100) | 17 (94) |
| Older adults (55–75 years) |  |  |  |  |  |  |  |  |  |  |  |
|  | N = |  | 8 | 8 |  | 8 | 8 |  | 8 | 8 | 12 |
| Age (yrs) | Mean |  | 62.8 | 63.1 |  | 59.1 | 60.3 |  | 59.8 | 61.5 | 62.3 |
|  | SD |  | 5.6 | 5.7 |  | 3.4 | 4.0 |  | 3.2 | 6.4 | 5.9 |
|  | range |  | (55–70) | (55–71) |  | (55–64) | (55–67) |  | (55–63) | (55–74) | (55–73) |
| Male | n (%) |  | 5 (63) | 4 (50) |  | 5 (63) | 5 (63) |  | 2 (25) | 4 (50) | 3 (25) |
| Race | n (%) |  |  |  |  |  |  |  |  |  |  |
| White |  |  | 8 (100) | 8 (100) |  | 8 (100) | 8 (100) |  | 8 (100) | 8 (100) | 12 (100) |
| Ethnicity | n (%) |  |  |  |  |  |  |  |  |  |  |
| Hispanic or Latino |  |  | - | 1 (13) |  | - | - |  | - | 1 (13) | 1 (8) |
| Not Hispanic or Latino |  |  | 8 (100) | 7 (88) |  | 8 (100) | 8 (100) |  | 8 (100) | 7 (88) | 11 (92) |

**Figure 1.**
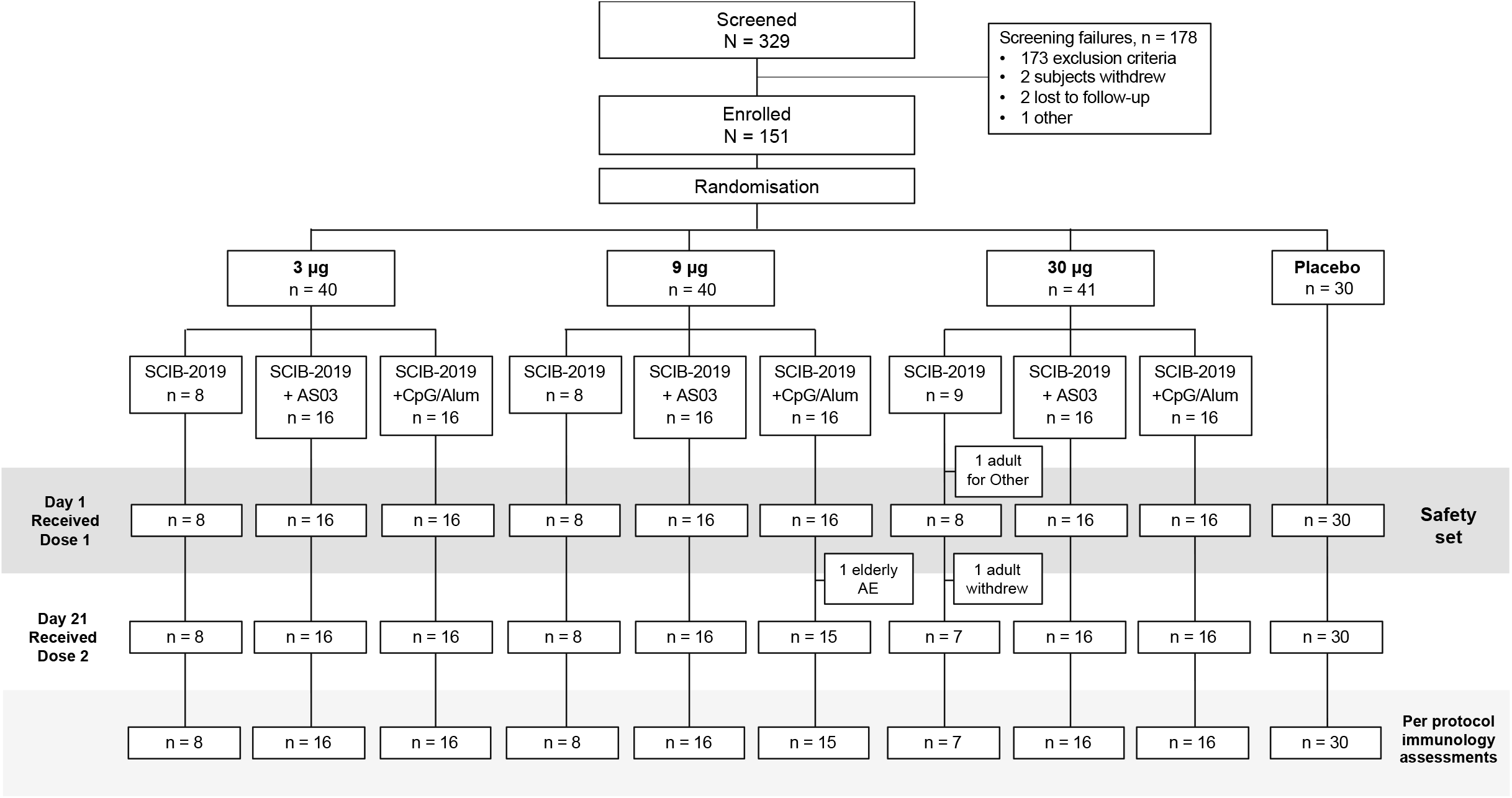
Study flow chart (all ages combined)

### Safety assessments

There were no deaths or hospitalisations during the study, and only two SAEs both in older adults. One older adult was diagnosed with cellulitis following a cat bite but completed the study, while another had hyponatraemia after receiving one dose in the 9 μg CpG/Alum group and was withdrawn from the study (**figure 1**). The participant was subsequently found to have a pituitary adenoma, which is a known to potentially cause hyponatremia [12–14]. Neither event was considered to be associated with vaccination. One adult decided to withdraw from the 30 μg SCB-2019 group for personal reasons before receiving their second vaccination. All other participants completed at least 21 days of follow-up after dose 2 and up to Day 50.

The non-adjuvanted trimeric-protein, SCB-2019, was generally well tolerated in terms of solicited local AEs with only one report of mild pain after a first 3 μg dose and none after second doses (**figure 2**). Formulations with AS03 resulted in approximately 50% of participants having local AEs after the first vaccination, consisting almost exclusively of transient Grade 1 or 2 injection site pain, cases of redness and swelling being relatively infrequent. The frequency and severity of local AEs tended to increase after the second dose, including one case of Grade 3 pain after a 9 μg dose of SCB-2019+AS03. When formulated with CpG/Alum, there was a SCB-2019 dose-dependent induction of Grade 1 local AEs, reported by 5 of 16 (44%) participants after the first dose and second doses of 30 μg SCB-2019+CpG/Alum. One Grade 3 case of pain was reported after as second dose of 9 μg SCB-2019+CpG/Alum. All local AEs were transient and resolved within the reporting period. Young adults reported local AEs more frequently (39% of all dose levels combined) than the older adults (21%) after the first dose, but incidence rates were similar in the two age groups, 35% and 34%, respectively, after the second dose.

**Figure 2.**
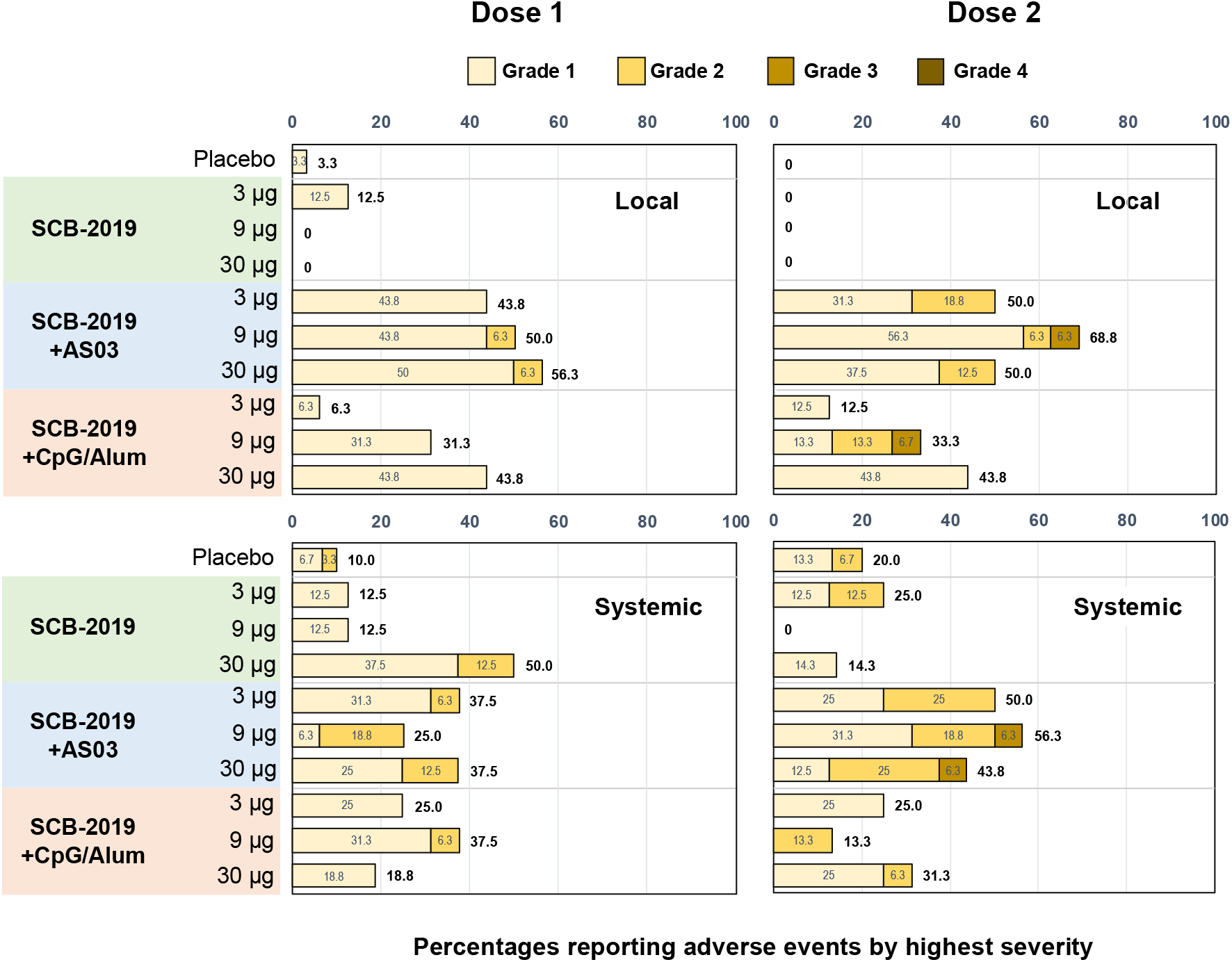
Incidence and severity of solicited local and systemic adverse events (all ages combined). There were no Grade 4 AEs reported.

After the first dose of plain SCB-2019 solicited systemic AEs were infrequent and similar to placebo in the 3 μg and 9 μg groups, but 50% of the 30 μg group reported Grade 1 or 2 AEs. These rates were lower after the second doses (**figure 2**). In contrast, when formulated with AS03 the frequency of systemic AEs was higher and not dose-dependent, reported by 25– 38% per group after the first dose and 44–56% after the second dose with a concomitant increase in the proportion described as Grade 2. Two participants, one each in the 9 μg and 30 μg groups, reported Grade 3 fatigue and myalgia. The most frequently reported systemic adverse events were headache, fatigue, and myalgia, with six reports of fever, all Grade 1 or 2 after the second dose of SCB-2019+AS03. Systemic AE rates in those who received SCB-2019+CpG/Alum were similar to the SCB-2019+AS03 group after the first dose, but there was no consistent trend to increase frequency or severity after the second dose. As with local AEs, the frequency of reported systemic AEs was lower in older adults after their first dose (17%) than the younger adults (38%), and overall rates were similar after second doses, 30% in older and 34% in younger adults, respectively. None of the participants took prophylactic paracetamol or nonsteroidal anti-inflammatory drugs.

### Unsolicited adverse events

Unsolicited adverse events reported over the 50-day study period mainly consisted of cases of Grade 1 or 2 headache or gastrointestinal disorders (nausea, abdominal pain) reported after the solicited reporting periods. There were no cases of unsolicited AEs causally related to vaccination and no trend for association with any particular groups. No consistent trends or clinically significant laboratory safety abnormalities were observed in any group at any timepoint.

No episodes of COVID-19 episodes or cases of SARS-CoV-2 infection were reported in the study. No adverse events of special interest, including potential immune-mediated diseases, were observed.

### Immunogenicity assessments - Anti-SCB-2019 IgG antibodies

Anti-SCB-2019 IgG antibodies did not increase after the first dose of unadjuvanted SCB-2019 by Day 22, irrespective of dose level (**figure 3**). By Day 50, three SCB-2019 recipients seroconverted, 1 of 8 (12·5%) in the 3 μg and 2 of 7 (28·6%) in the 30 μg group (**table 2**), although GMTs were low. In both adjuvanted cohorts there were SCB-2019 dose-dependent IgG responses evident after a single dose in both age groups (**figure 3**). All participants at each dose level of SCB-2019+AS03 seroconverted by Day 36 (**table 2**). Following a second dose of SCB-2019+AS03 there were marked increases in GMTs, to higher levels than those observed in the convalescent sera (GMT = 666 EC_50_ [95% CI: 272–1628], n = 21) with a range of GMTs from 2510 to 4452 across the SCB-2019+AS03 dose levels in young adults, 1567 to 3625 in older adults (**figure 3**). Antibody titres persisted at high levels at day 50.

**Table 2.** Antibody seroconversion rates in young and elderly adult groups combined.

| Seroconversion rate |  |  | 3 µg SCB-2019 |  |  | 9 µg SCB-2019 |  |  | 30 µg SCB-2019 |  |  |
| --- | --- | --- | --- | --- | --- | --- | --- | --- | --- | --- | --- |
| (SCR) |  | Placebo | Plain | AS03 | CpG/Alum | Plain | AS03 | CpG/Alum | Plain | AS03 | CpG/Alum |
| Anti-SCB-2019 IgG antibodies |  |  |  |  |  |  |  |  |  |  |  |
| Day 22 | n = | 30 | 8 | 16 | 16 | 8 | 16 | 15 | 7 | 16 | 16 |
|  | SCR, n (%) | 0 | 0 | 7 (43.8) | 0 | 0 | 9 (56.3) | 2 (13.3) | 0 | 14 (87.5) | 5 (31.3) |
| Day 36 | n = | 30 | 8 | 16 | 16 | 8 | 16 | 15 | 7 | 16 | 15 |
|  | SCR, n (%) | 0 | 1 (12.5) | 16 (100) | 13 (81.3) | 0 | 16 (100) | 14 (93.3) | 1 (14.3) | 16 (100) | 14 (93.3) |
| Day 50 | n = | 29 | 8 | 16 | 16 | 8 | 16 | 15 | 7 | 16 | 15 |
|  | SCR, n (%) | 0 | 1 (12.5) | 15 (93.8) | 14 (87.5) | 0 | 16 (100) | 14 (93.3) | 2 (28.6) | 16 (100) | 14 (93.3) |
| ACE2-receptor competitive-binding IgG antibodies |  |  |  |  |  |  |  |  |  |  |  |
| Day 22 | n = | 30 | 8 | 16 | 16 | 8 | 16 | 15 | 7 | 16 | 16 |
|  | SCR, n (%) | 0 | 0 | 4 (25.0) | 0 | 0 | 2 (12.5) | 2 (13.3) | 0 | 6 (37.5) | 1 (6.3) |
| Day 36 | n = | 30 | 8 | 16 | 16 | 8 | 16 | 15 | 7 | 16 | 15 |
|  | SCR, n (%) | 0 | 0 | 15 (93.8) | 8 (50.0) | 0 | 16 (100) | 12 (80.0) | 1 (14.3) | 16 (100) | 14 (93.3) |
| Day 50 | n = | 29 | 8 | 16 | 16 | 8 | 16 | 15 | 7 | 16 | 15 |
|  | SCR, n (%) | 0 | 0 | 15 (93.8) | 8 (50.0) | 0 | 16 (100) | 12 (80.0) | 0 | 16 (100) | 14 (93.3) |
| Wild-type SARS-CoV-2 neutralising antibodies |  |  |  |  |  |  |  |  |  |  |  |
| Day 22 | n = | 30 | 8 | 16 | 16 | 8 | 16 | 15 | 7 | 16 | 16 |
|  | SCR, n (%) | 0 | 0 | 2 (12.5) | 0 | 0 | 7 (43.8) | 2 (13.3) | 0 | 7 (43.8) | 4 (25.0) |
| Day 36 | n = | 30 | 8 | 16 | 16 | 8 | 16 | 15 | 7 | 16 | 15 |
|  | SCR, n (%) | 0 | 0 | 15 (93.8) | 14 (87.5) | 0 | 16 (100) | 13 (86.7) | 1 (14.3) | 16 (100) | 14 (93.3) |
| Day 50 | n = | 28 | 8 | 16 | 16 | 8 | 16 | 15 | 7 | 16 | 15 |
|  | SCR, n (%) | 0 | 0 | 15 (93.8) | 12 (75.0) | 0 | 16 (100) | 13 (86.7) | 0 | 16 (100) | 14 (93.3) |

**Figure 3.**
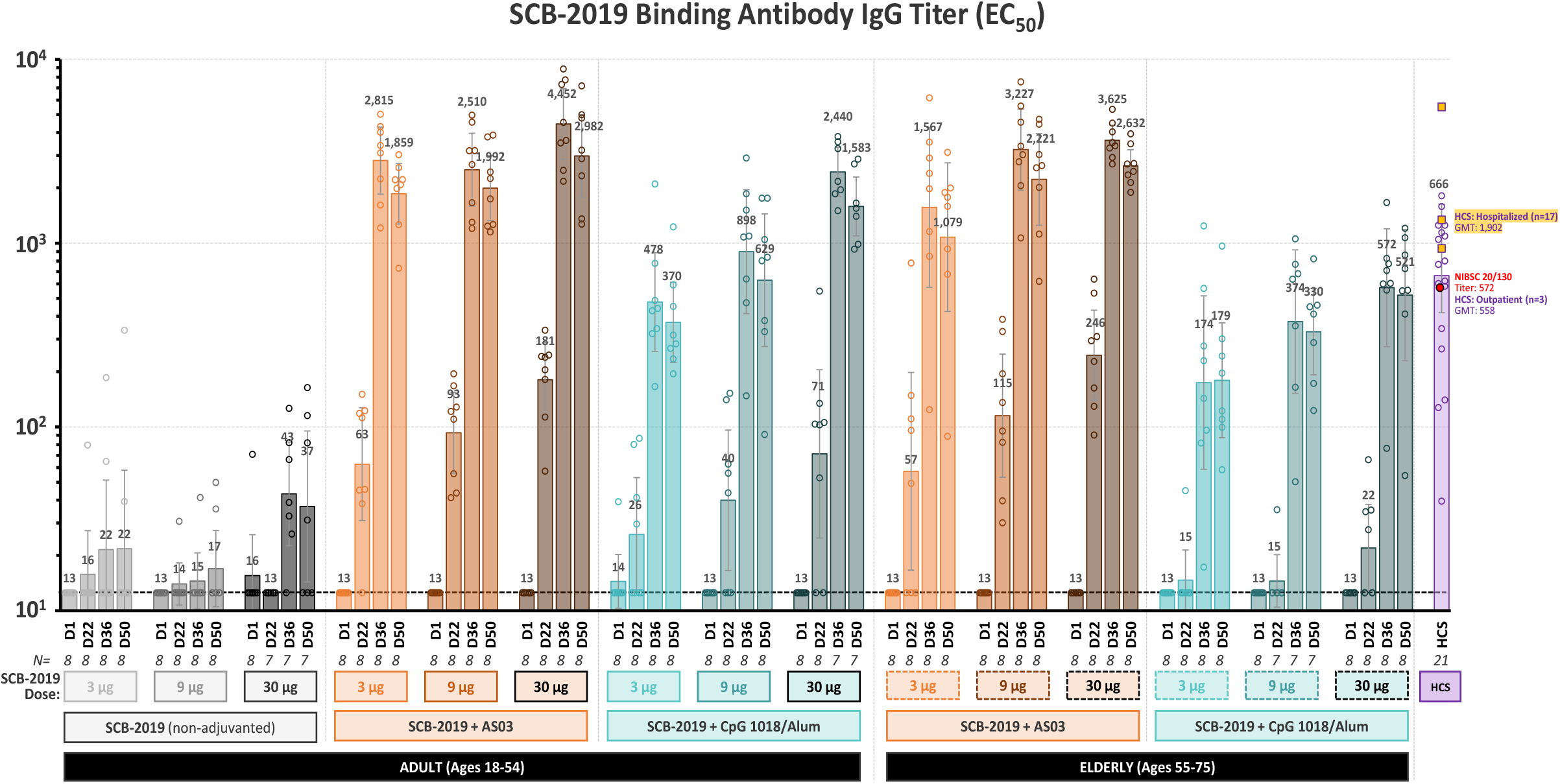
SCB-2019 IgG antibody titres. IgG antibodies binding to SCB-2019 in the different study groups and the COVID-19 human convalescent sera measured by ELISA (EC_50_). Bars show geometric mean titre (GMT) values per group with 95% confidence intervals at Days 1, 22, 36 and 50. Circles represent values for individual subjects.

Small dose-dependent IgG responses against SCB-2019 in the 3 μg, 9 μg and 30 μg SCB-2019+CpG/Alum groups at Day 22 after a single dose in young adults markedly increased after the second dose with GMTs of 478, 898 and 2440, respectively, on Day 36. GMTs were lower in the equivalent older adult groups; 174, 374 and 572 at Day 36 (**figure 3**). High GMTs were maintained to Day 50, when seroconversion rates were 87·5–93·8% in both age groups combined across doses (**table 2**).

### Immunogenicity assessments - ACE2-competitive binding ELISA

The profile of ACE2 receptor-competitive binding antibody responses was similar those observed as SCB-2019 IgG (**figure 4**). There was little or no response to unadjuvanted SCB-2019 but robust responses to the first and second doses of SCB-2019+AS03 for all dose levels which achieved similar GMTs in both age groups at Day 36, ranging from 435–754 in young adults and 288–688 in the older adults across all three dose levels. Seroconversion rates in all ages were 93·8%, 100% and 100% for the 3 μg, 9 μg and 30 μg dose levels All groups had GMTs that were higher than those observed in convalescent sera; 144 (95% CI: 54–386], n = 21), and these remained higher than convalescent sera at Day 50. Following SCB-2019+CpG/Alum there were dose-dependent responses in young adults which were higher than in the older adults, and only matched the levels in convalescent sera with 9 μg and 30 μg doses of SCB-2019+CpG/Alum in the young adult groups, who had seroconversion rates of 50%, 80% and 93·3% after two doses (**table 2**).

**Figure 4.**
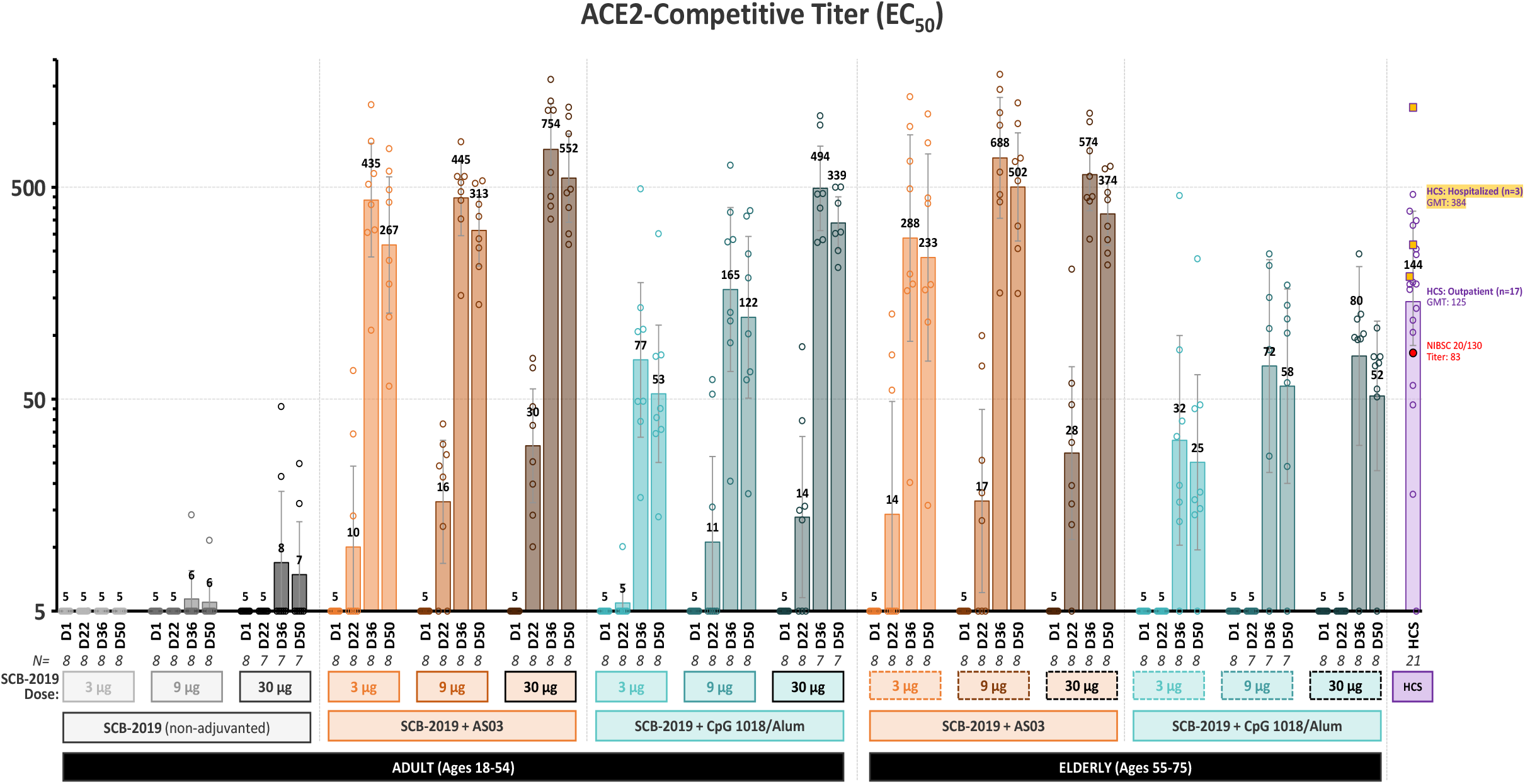
ACE2-competitive titres in the different study groups and the COVID-19 human convalescent sera measured by ELISA (EC_50_). Bars show geometric mean titre (GMT) values per group with 95% confidence intervals at Days 1, 22, 36 and 50. Circles represent values for individual subjects.

### Immunogenicity assessments – SARS-CoV-2 neutralising activity

Serum neutralising activity of wild-type SARS-CoV-2 virus showed a similar pattern of responses to SCB-2019-binding IgG antibodies (**figure 5**). No increase in neutralising activity was observed with unadjuvanted SCB-2019, with only 1 of 7 (14%) participants responding by Day 36 in the 30 μg group (**table 2**). Following the first dose of SCB-2019+AS03, 16 of 48 (33%) recipients across doses had seroconverted, increasing to 47 of 48 (98%) by Day 36, after the second dose. This increase in neutralising antibodies was SCB-2019 dose-dependent, shown by the geometric means of 1280, 1810 and 3948 MN_50_ in the 3 μg, 9 μg and 30 μg groups, respectively. Importantly the range of MN_50_ GMTs seen in older adult groups (1076–3320) were similar to those in the young adult groups for all dose levels in both age groups and were higher than convalescent sera (MN_50_ GMT 717; 95% CI 213-2417, n = 21). There was some decline in GMTs, but high levels of neutralising antibodies persisted to Day 50 in both age groups (**Figure 5**).

**Figure 5.**
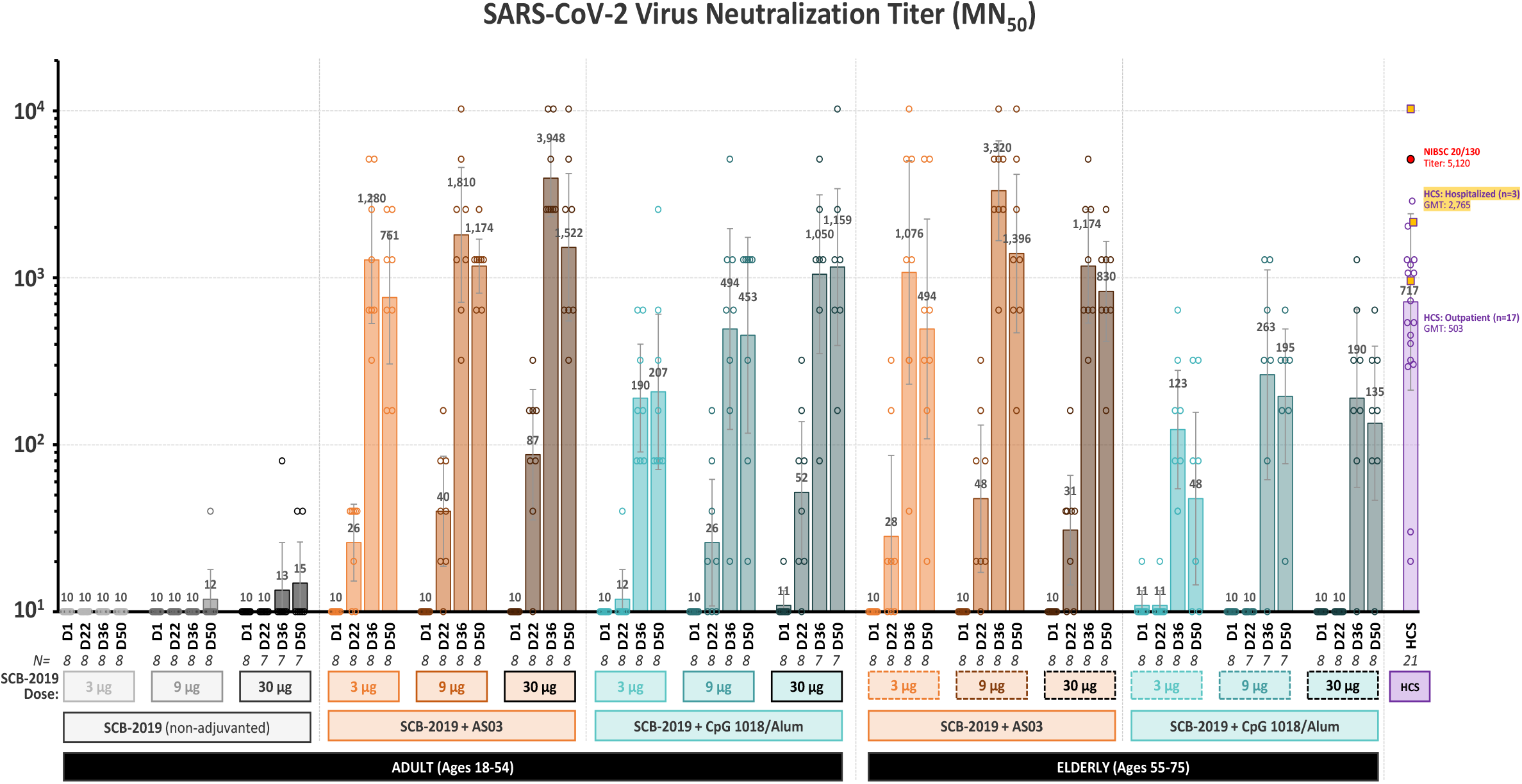
Wild-type SARS-CoV-2 virus neutralisation titres in the different study groups and the COVID-19 human convalescent sera measured by microneutralization based on CPE (MN_50_). Bars show geometric mean titre (GMT) values per group with 95% confidence intervals at Days 1, 22, 36 and 50. Circles represent values for individual subjects.

Dose-dependent increases in neutralising activity were also observed in the SCB-2019+CpG/Alum groups, but these responses were lower in magnitude than the AS03 groups as illustrated in **figure 5**. In the older adult groups the range of MN_50_ GMTs (123–263) appeared lower than in the convalescent sera. High titres were maintained up to Day 50, the last timepoint tested in this interim analysis.

### Relationship of different antibody assays

When the correlations between immune responses assessed by the three different assays were investigated in convalescent and vaccinee sera there were highly significant linear relationships between each of the three assays (*see Supplementary appendix pages 6–7*). The calculated Pearson correlation coefficients were R = 0·88, p<0·001 for ACE2-competitive vs. SCB-2019 binding IgG antibodies, R = 0·70, p<0·001 for SCB-2019 binding IgG antibodies vs. VNT, and R = 0.67, p<0·001 for VNT vs. ACE2-competitive binding antibodies. Further, the calculated ratios of neutralising antibodies to SCB-2019 binding IgG antibodies in vaccinee sera obtained at Day 36 and 50, following the two doses of vaccines, fell in the same range for SCB-2019+AS03 and SCB-2019+CpG/Alum and convalescent sera (*see Supplementary appendix page 8*).

### Cell-mediated immune (CMI) responses

Assessment of Th1-biased cell-mediated immune responses specific to the SARS-CoV-2 spike protein were observed in both adjuvanted vaccine groups with increases in interferon-γ and/or IL-2 positive CD4+ T-cells after the first dose which further increased after the second dose (*see Supplementary appendix page 9*). There were no CMI responses with unadjuvanted SCB-2019, and no observable increases in Th2 (IL-4 or IL-5 positive CD4+ cells) or Th17 (IL-17 positive) cellular immune responses in any group.

## DISCUSSION

The primary objective of this study was to assess the safety and reactogenicity of SCB-2019 when administered alone or as one of two adjuvanted formulations with AS03 or CpG and Alum. All formulations appeared safe as there were no vaccine-related SAEs or study withdrawals. Plain SCB-2019 was well tolerated and when administered in combination with either adjuvant, demonstrated acceptable reactogenicity with few Grade 3 solicited adverse events. Higher reactogenicity was observed with AS03, unaffected by the dose level of SCB-2019, which consisted of mainly transient Grade 1 or 2 adverse events that all resolved spontaneously without intervention. No prophylactic paracetamol was used in this study and would not appear necessary for general use. When the age of the participants was considered there was no overall impact on the safety or reactogenicity. Although the older adults (55–75 years) displayed fewer local and systemic adverse events than younger adults (18–54 years) after the first dose, incidence rates of solicited adverse events were similar in both age groups after the second dose. The use of AS03 in pandemic H5N1 influenza vaccines allowed a demonstration of its general safety [15], while large trials of the same vaccine revealed a higher local reactogenicity than we observed with injection pain in 89% of 18–64 year-olds [16]. Overall, this reactogenicity profile compares favourably with those of the mRNA SARS-CoV-2 vaccines which had incidence rates of local pain rates approaching or reaching 100% in adults [17,18]. The rates of solicited AEs in the CpG/Alum-adjuvanted vaccine groups were lower and consistent with licensed CpG-adjuvanted vaccine [19,20].

SCB-2019 alone at the tested dose levels was poorly immunogenic but when combined with ether adjuvant system it elicited robust increases in functional immune responses detected as SARS-CoV-2 virus neutralising activity that correlated well with IgG antibodies against SCB-2019 or ACE2 competitive binding antibodies. Neutralising responses were already observed after the first dose in the higher dose AS03 groups. Highest responses were observed with AS03 and after completion of the two dose series GMTs rapidly peaked at Day 36 at levels that were higher than those observed in convalescent sera from patients hospitalised with COVID-19 and the NIBSC standard sample. These high levels persisted until the end of this interim analysis as Day 50. There was little meaningful difference between the immune responses to SCB-2019+AS03 between the young and older adults. When adjuvanted with CpG and Alum, the immune responses were lower than with AS03 and SCB-2019 dose-dependent. Further, the response to SCB-2019+CpG/Alum were lower in the older age group. Further investigation of the cellular immune responses showed increases in Th1-polarised responses after both first and second doses for both AS03-adjuvanted and CpG/Alum-adjuvanted SCB-2019. CD4+ T-cell responses have been suggested to complement humoral antibody responses in overcoming SARS-CoV-2 infection [19].

As a strong correlation between the neutralising activity and ELISA IgG antibody responses to S protein and RBD site has been seen in convalescent sera from PCR-confirmed COVID-19 patients [21] we investigated these ratios in our assays. We confirmed strong correlations between neutralising activity and IgG measured in either the SCB-2019 or ACE2-receptor assays. This is an important aspect of the immune response as it has been suggested that low neutralising/binding antibody ratios could contribute to an increased risk of antibody-enhanced disease [22]. Both adjuvanted formulations have been shown to be protective in preclinical non-human primate and rodent animal models, but AS03 formulations appeared to induce superior humoral immunogenicity and we observed an apparent lack of age-effect on the response. Although the AS03 formulation had higher reactogenicity compared with CpG/Alum, the severity appeared to be lower than to the mRNA and some vector-derived SARS-CoV-2 vaccines [23–25] and mainly consisted of transient mild to moderate local and systemic adverse events.

This study is limited in being relatively small with only 8 participants per vaccine formulation and age group, and in seronegative participants only, but overall it confirms the apparent safety and general tolerability of the different formulations. The small size also hampers observation of any true impact of age on tolerability or immunogenicity. Further, the immune responses have only been assessed up to Day 50, approximately four weeks after the second vaccination, so we have no data on persistence of the response or waning of the immunity. These limitations are being addressed in the extension of the study. While we have compared the immune responses elicited by vaccination with those measured in the convalescent sera of patients with PCR-confirmed COVID-19 infection, in the absence of an established serologic correlate of protection we cannot extrapolate our data to infer protection.

In conclusion we have demonstrated safety, good tolerability and high neutralising immune responses with a Th1-biased cellular immune response that supports the further development of both candidate vaccines including assessment of the protective efficacy.

## Data Availability

The datasets, including the redacted study protocol, redacted statistical analysis plan, and individual participants data supporting the results reported in this article, will be available three months from initial request, to researchers who provide a methodologically sound proposal. The data will be provided after its de-identification, in compliance with applicable privacy laws, data protection and requirements for consent and anonymisation.

## ACKNOWLEDGEMENTS

We are grateful to all the volunteers and the study staff at Nedlands, Western Australia. We wish to thank GSK and Dynavax for providing the AS03 and CpG 1018 adjuvants, respectively, and the Melbourne Health and VIDRL (Victorian Infectious Diseases Reference Laboratory) for providing a sample of SARS-CoV-2 for use in the virus neutralisation assays. The authors are grateful to Professor Jim Buttery and the SPEAC, and the Scientific Advisory Board (Donna Ambrosino, Sue Ann Costa Clemens, Pierre Desmons, Sam Liao, Michael Pfleiderer, Antoinette Quinsaat, Frank Rockhold, David Salisbury, George Siber, Nelson Teich, Anh Wartel, and Nicholas Jackson) for helpful expert advice and support. We thank Keith Veitch (keithveitch communications, Amsterdam, the Netherlands) for drafting and editorial management of the manuscript.

## FINANCIAL DECLARATION

This work was supported by grants from Coalition for Epidemic Preparedness Innovations (CEPI).

## DISCLAIMER

Any opinions, findings, and conclusions expressed in this material are those of the authors.

## CONTRIBUTORS

MD, JL, RC, PR, BM, BH, PL, IS and HHH designed the study, data collection was by LH, and data analysis support by PL. Interpretation and writing of the manuscript was by all authors led by RC and a medical writer. All authors approved the submission for publication.

## SARS-CoV-2 Virus Neutralization Titer (MN_50_)

### Supplementary Appendix

### Definition of a Serious Adverse Event (SAE)

An SAE is any adverse event that:

- Results in death
- Is life-threatening
- Requires inpatient hospitalisation or prolongation of existing hospitalisation
- Results in persistent disability/incapacity
- Is a medically important event in the opinion of the investigator

### Definition of an Adverse Event of Special Interest (AESI)

An AESI is a potential immune-mediated disease including autoimmune disease and other inflammatory and/or neurologic disorder of interest which may or may not have an autoimmune aetiology.

#### Grades for Solicited Local Reactions

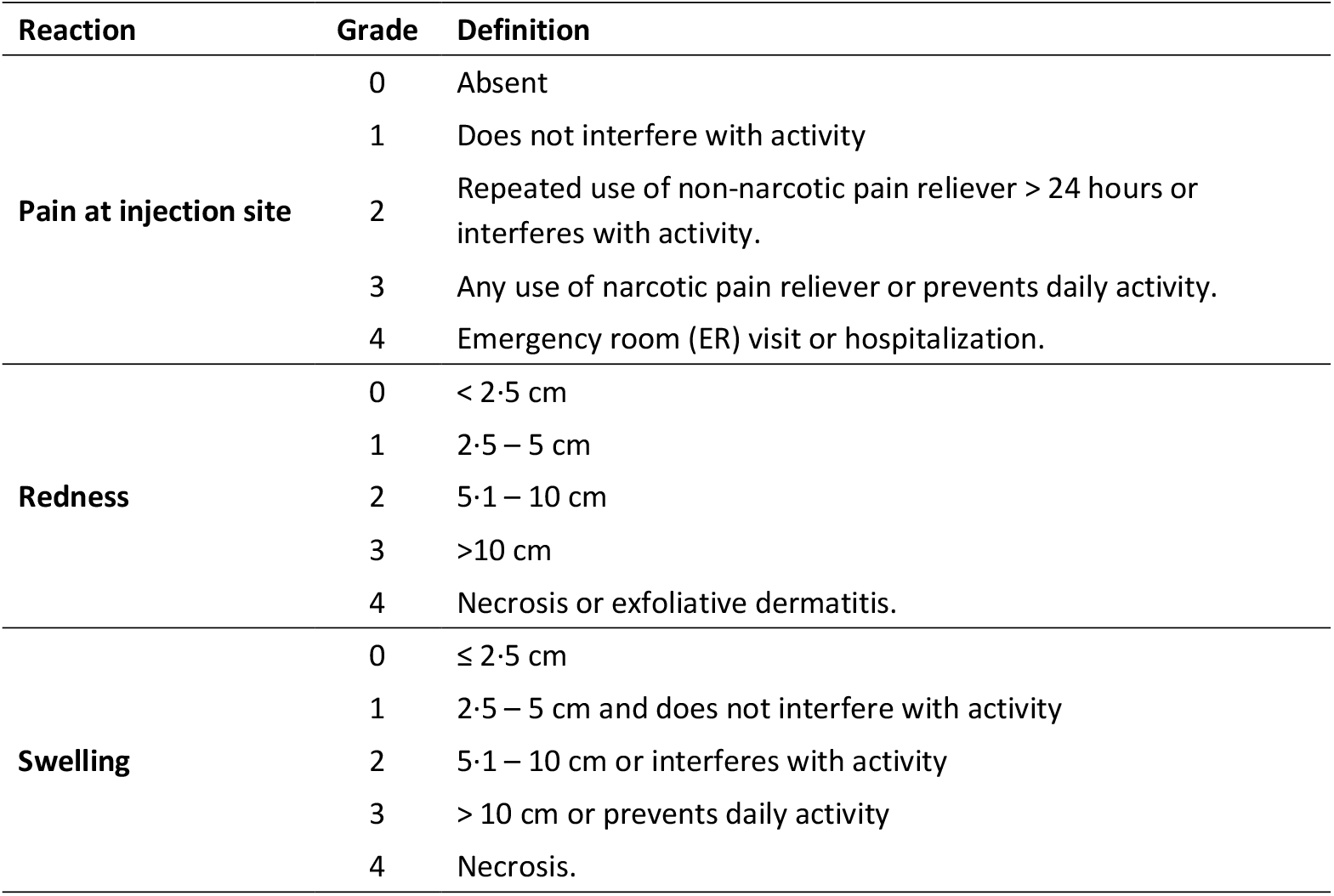

| Reaction | Grade | Definition |
| --- | --- | --- |
| **Pain at injection site** | 0 | Absent |
|  | 1 | Does not interfere with activity |
|  | 2 | Repeated use of non-narcotic pain reliever > 24 hours or interferes with activity. |
|  | 3 | Any use of narcotic pain reliever or prevents daily activity. |
|  | 4 | Emergency room (ER) visit or hospitalization. |
| **Redness** | 0 | < 2.5 cm |
|  | 1 | 2.5 – 5 cm |
|  | 2 | 5.1 – 10 cm |
|  | 3 | >10 cm |
|  | 4 | Necrosis or exfoliative dermatitis. |
| **Swelling** | 0 | ≤ 2.5 cm |
|  | 1 | 2.5 – 5 cm and does not interfere with activity |
|  | 2 | 5.1 – 10 cm or interferes with activity |
|  | 3 | > 10 cm or prevents daily activity |
|  | 4 | Necrosis. |

#### Grades for Solicited Systemic Adverse Events

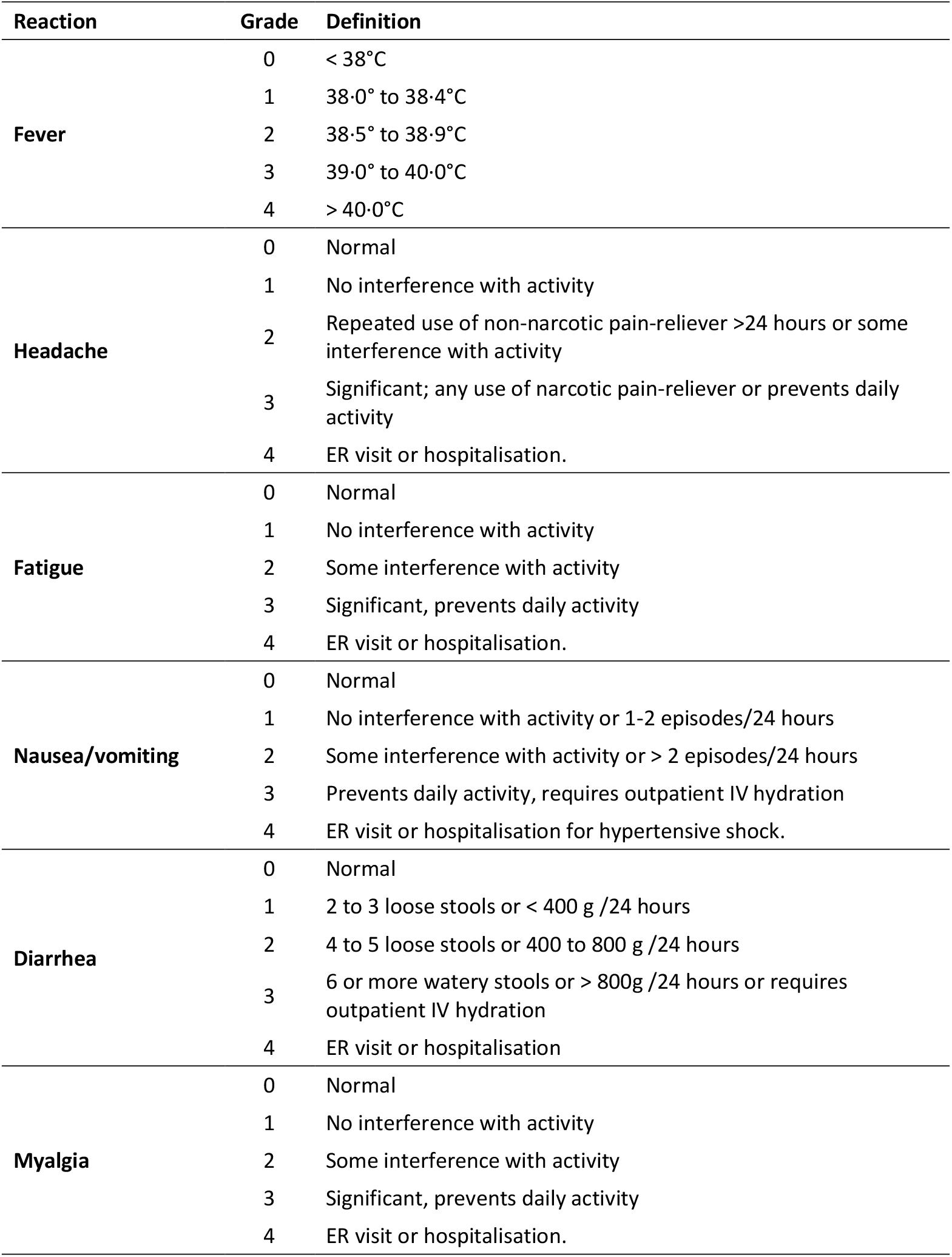

| **Reaction** | **Grade** | **Definition** |
| --- | --- | --- |
| **Fever** | 0 | < 38°C |
|  | 1 | 38.0° to 38.4°C |
|  | 2 | 38.5° to 38.9°C |
|  | 3 | 39.0° to 40.0°C |
|  | 4 | > 40.0°C |
| **Headache** | 0 | Normal |
|  | 1 | No interference with activity |
|  | 2 | Repeated use of non-narcotic pain-reliever >24 hours or some interference with activity |
|  | 3 | Significant; any use of narcotic pain-reliever or prevents daily activity |
|  | 4 | ER visit or hospitalisation. |
| **Fatigue** | 0 | Normal |
|  | 1 | No interference with activity |
|  | 2 | Some interference with activity |
|  | 3 | Significant, prevents daily activity |
|  | 4 | ER visit or hospitalisation. |
| **Nausea/vomiting** | 0 | Normal |
|  | 1 | No interference with activity or 1-2 episodes/24 hours |
|  | 2 | Some interference with activity or > 2 episodes/24 hours |
|  | 3 | Prevents daily activity, requires outpatient IV hydration |
|  | 4 | ER visit or hospitalisation for hypertensive shock. |
| **Diarrhea** | 0 | Normal |
|  | 1 | 2 to 3 loose stools or < 400 g /24 hours |
|  | 2 | 4 to 5 loose stools or 400 to 800 g /24 hours |
|  | 3 | 6 or more watery stools or > 800g /24 hours or requires outpatient IV hydration |
|  | 4 | ER visit or hospitalisation |
| **Myalgia** | 0 | Normal |
|  | 1 | No interference with activity |
|  | 2 | Some interference with activity |
|  | 3 | Significant, prevents daily activity |
|  | 4 | ER visit or hospitalisation. |

### IMMUNOLOGICAL METHODS

#### SCB-2019 binding ELISA

Maxisorp plates were coated with 1μg/ml SCB-2019 at 4°C overnight and blocked with 2% non-fat milk in PBS containing 0·05% Tween-20 (PBST). Eight two-fold serial dilutions of serum samples starting from a 1:25 initial dilution were added to the blocked and washed SCB-2019-coated plates and incubated for 1h at 37°C. Plates were then washed, incubated with HRP-conjugated anti-human IgG for 1 hr at 37°C, washed again and the colorimetric signals were developed using TMB substrate for 3 min before stopping the reaction with 1N sulfuric acid. Optical density (OD) was measured at 450/650 nm. The EC_50_ of each test sample was calculated using a non-linear four parameter regression curve using GraphPad Prism, v.6.0c.

#### ACE2-competitive binding ELISA

Maxisorp plates were coated with 1 μg/mL ACE2-Fc at 4°C overnight, blocked with 2% non-fat milk in PBS containing 0·05% Tween-20 (PBST). On dilution plates equal volumes of eight two-fold, serial-diluted sera starting from a 1:5 initial dilution were incubated with biotinylated SCB-2019 (200 ng/mL) for 30 min at ambient temperature. Vials of negative normal human serum and negative normal human serum spiked with 200 ng/ml biotinylated SCB-2019 were prepared as negative and positive controls, respectively. After incubation, mixtures and quadruplicate negative and positive controls were transferred to the blocked and washed ACE2-Fc coated plates and incubated for a further 1 hr at 37°C. The plates were then washed and incubated with HRP-conjugated streptavidin at 37°C for 1 h. Colorimetric signals were developed using TMB substrate until the OD650 of the positive control wells reached 0·7 when the reaction was stopped by addition of 1N sulfuric acid to the whole plate, and absorbance was promptly read at OD450/650nm. The 50% inhibition was calculated based on the negative and positive controls, and a non-linear four parameter regression curve was used to calculate the IC_50_ for each test sample with GraphPad Prism, v.6.0c.

#### SARS-CoV-2 wild-type microneutralisation (WT-MN)

Serum samples were heat inactivated for 30 minutes at 56°C and eleven two-fold serial dilutions of test samples were prepared in a separate dilution plate. Sera were mixed with an equal volume of SARS-CoV-2, hCoV-19/Australia/VIC01/2020 (GenBank MT007544.1), and incubated for 1 hr at 37°C, 5% CO_2_. The virus/serum mixtures (200 TCID_50_ units/well) were then transferred in duplicate to sub-confluent Vero E6 cell monolayer plates, pre-seeded 24 hours beforehand in 96 well plates at 1·5 ⨯ 10^4^ cells/well. Plates were incubated for 3 days at 37°C, 5% CO_2_. The residual non-neutralised virus was detected via cytopathic effect (CPE) by microscopic scoring. The neutralisation titre is expressed as the reciprocal of the highest dilution at which 50% of the replicate wells were protected from infection (MN_50_). If at least 50% protection was not observed for an individual serum sample at any dilution, the MN_50_ titre was recorded as <20.

### Correlation between different antibody response measurements in human convalescent sera

Shown are scatter plots of antibody titres comparing (A) ACE2-competitive titres versus anti-SCB-2019 IgG titres, (B) SARS-CoV-2 virus neutralisation titres versus anti-SCB-2019 IgG titres, and (C) SARS-CoV-2 virus neutralisation titres versus ACE2-competitive titres. This includes data for 21 sera samples (panel of 20 convalescent sera + NIBSC 20/130 preliminary reference sera).

R = Pearson correlation coefficient (GraphPad Prism, v.6.0c).

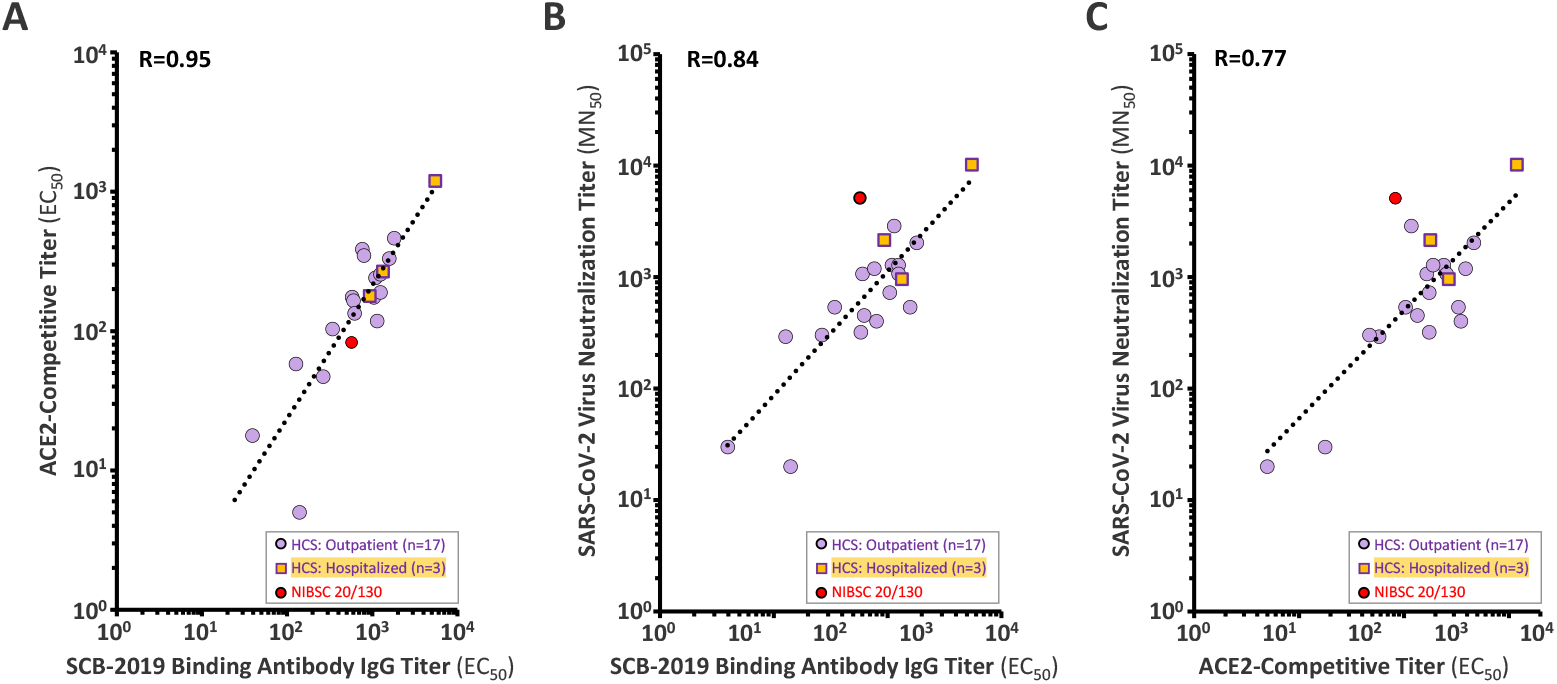

### Correlation between different antibody response measurements in vaccinees

Shown are scatter plots of antibody titres comparing (A) ACE2-competitive titres versus anti-SCB-2019 IgG titres, (B) SARS-CoV-2 virus neutralisation titres versus anti-SCB-2019 IgG titres, and (C) SARS-CoV-2 virus neutralisation titres versus ACE2-competitive titres. This includes data for all samples with a detectable SARS-CoV-2 virus neutralisation titre (MN_50_ ≥ 20); 126 observations in SCB-2019+AS03 vaccine groups, and 96 observations in SCB-2019+CpG/Alum vaccine groups.

R = Pearson correlation coefficient (GraphPad Prism, v.6.0c).

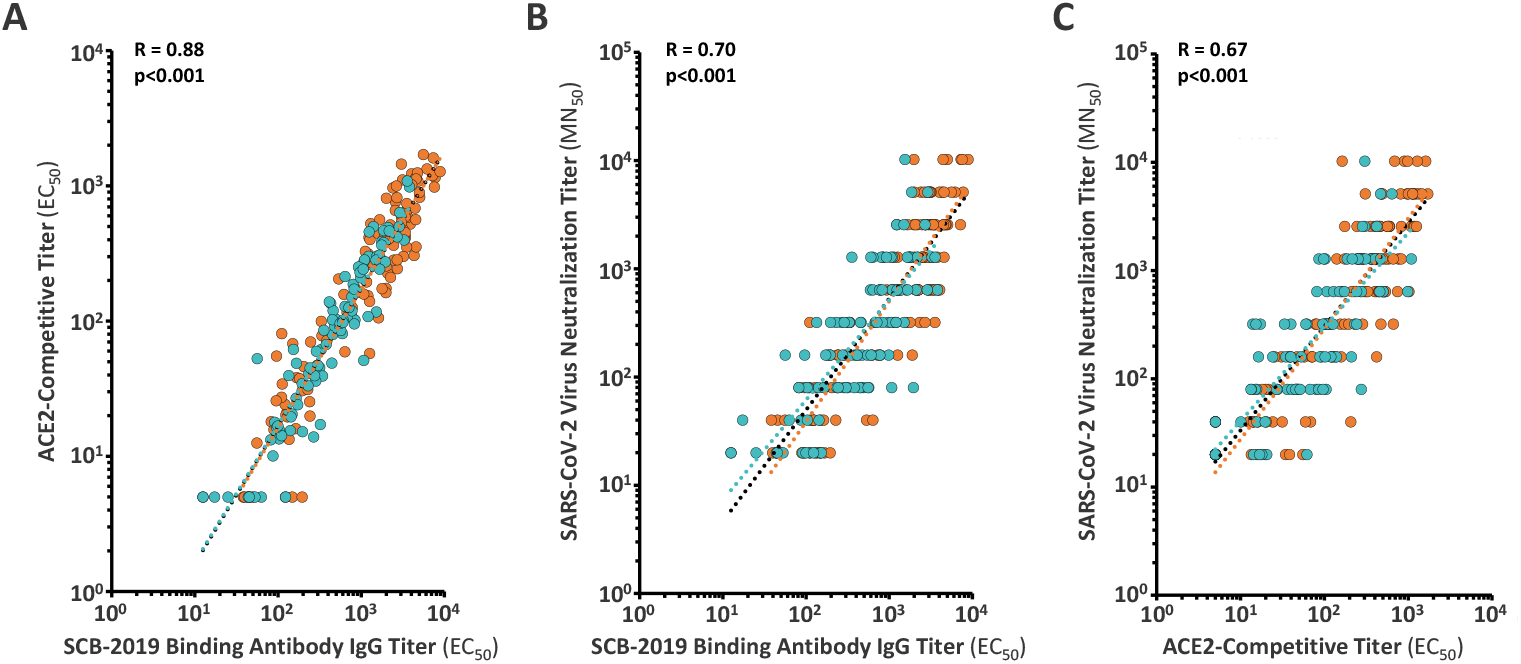

### Ratio of neutralising antibodies (VNT) to SCB-2019 binding antibodies (ELISA)

Shown are the ratios of wild-type SARS-CoV-2 virus neutralisation titres (MN_50_) to SCB-2019 binding antibody IgG titres (EC_50_). Bars show geometric mean values for each group ± 95% confidence intervals (CI). Dots represent values from individual subjects. Yellow highlighted range represents min-max range (0·14–2·5) of ratios in convalescent sera (excluding NIBSC 20/130).

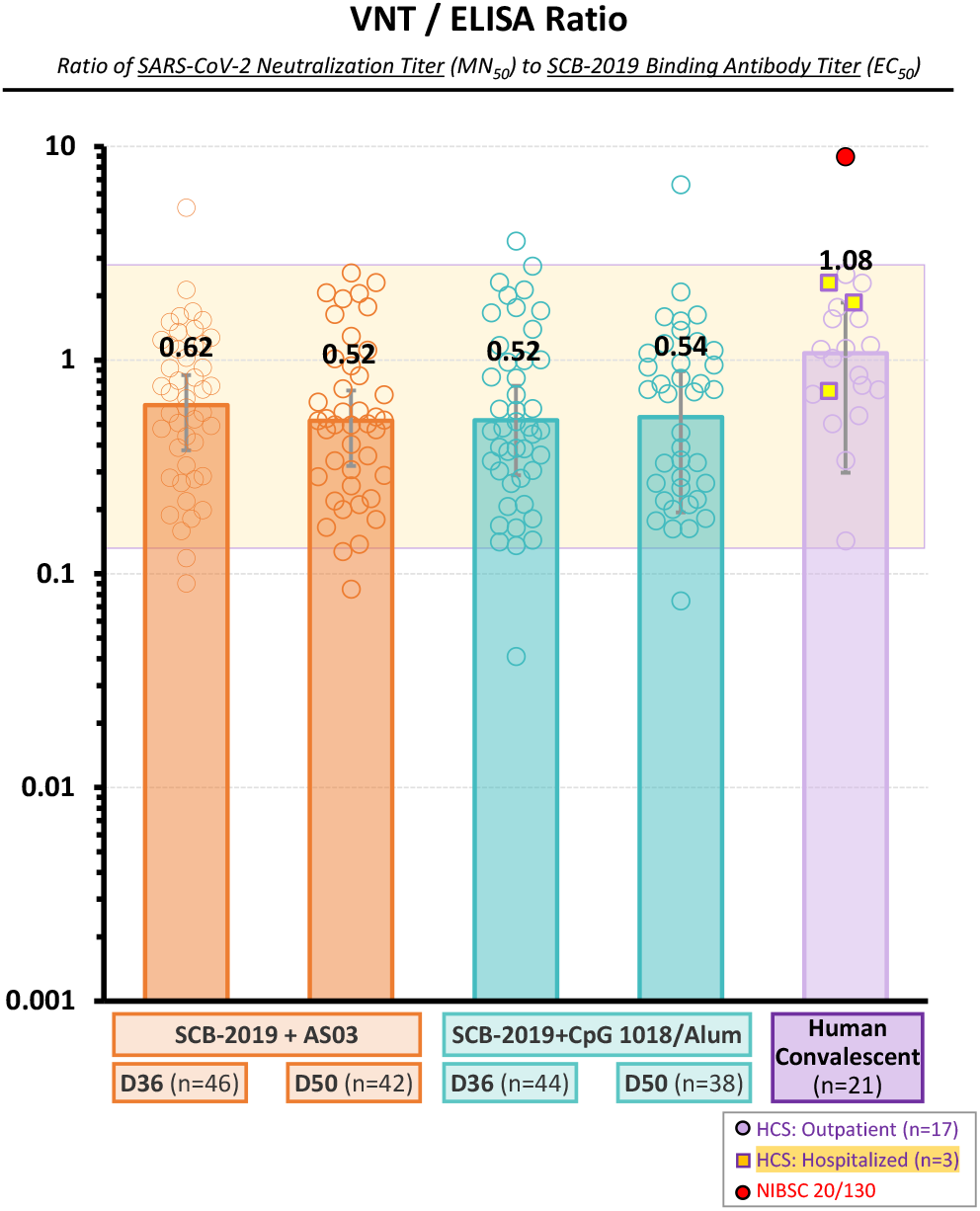

### SARS-CoV-2 CD4+ T-cell responses

Frequencies of SARS-CoV-2 Spike (S) protein-specific CD4+ cells expressing **(A)** Th1 cytokines (IFN-*γ* and/or IL-2), **(B)** Th2 cytokines (IL-4 and/or IL-5), or **(C)** Th17 cytokine (IL-17).. Bars show median values per group, and error bars represent interquartile range (IQR). Dots represent values for individual subjects.

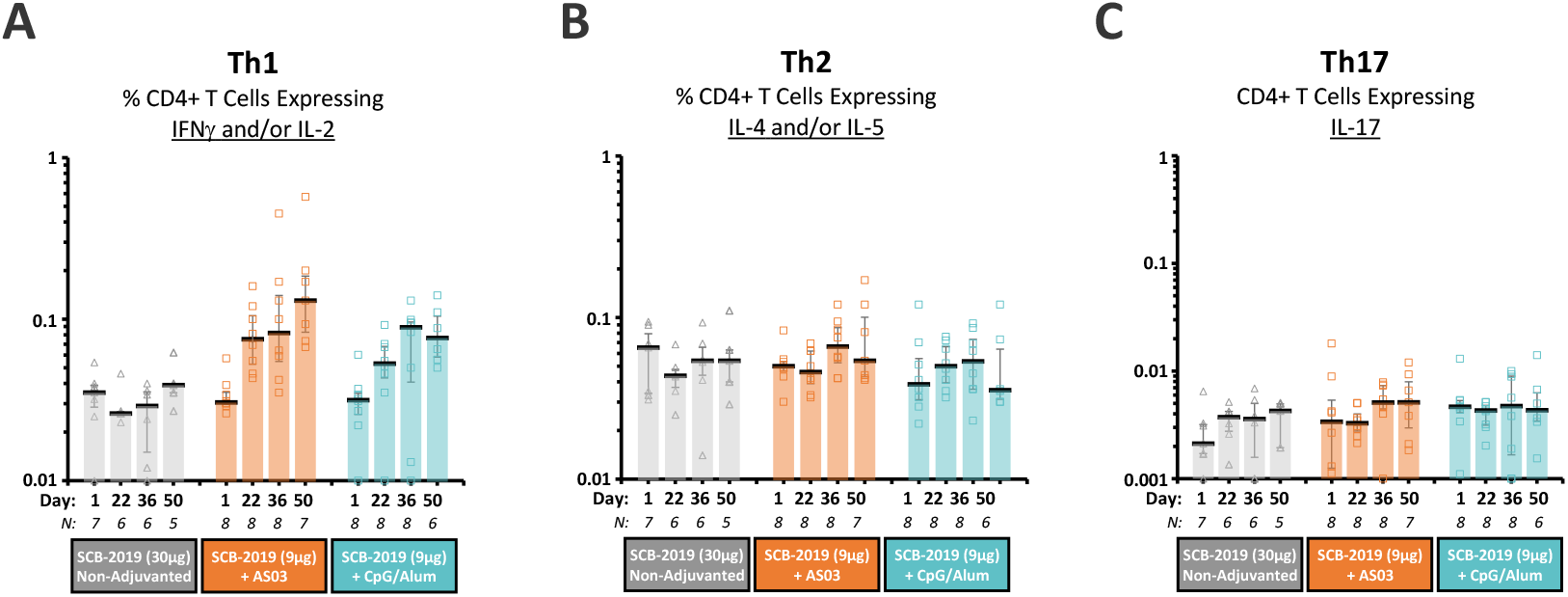

## REFERENCES

1. Johns Hopkins University of Medicine, Coronavirus Resource Center (November 11, 2020) https://coronavirus.jhu.edu/map.html.

2. Tan E, Song J, Deane AM, Plummer MP. Global impact of COVID-19 infection requiring admission to the intensive care unit: a systematic review and meta-analysis. Chest. 2020 Oct 15:S0012-3692(20)34906-0. doi: 10.1016/j.chest.2020.10.014.

3. WHO. DRAFT landscape of COVID-19 candidate vaccines–3 November 2020. https://www.who.int/publications/m/item/draft-landscape-of-covid-19-candidate-vaccines (accessed Nov 11, 2020).

4. Huang Y, Yang C, Xu X-f, Xu W, Liu S-w. Structural and functional properties of SARS-CoV-2 spike protein: potential antivirus drug development for COVID-19. Acta Pharmacologica Sinica 2020;41:1141–1149.

5. Shang J, Wan Y, Luo C, et al. Cell entry mechanisms of SARS-CoV-2. PNAS 2020;117:11727–11734.

6. Belouzard S, Millet JK, Licitra, BN, Whittaker GR. Mechanisms of coronavirus cell entry mediated by the viral spike protein. Viruses 2012;4:1011–1033.

7. Florindo HE, Kleiner R, Vaskovich-Koubi D, et al. Immune-mediated approaches against COVID-19. Nature Nanotechnology 2020;15:630–645.

8. Poland GA, Ovsyannikova IG, Kennedy RB. SARS-CoV-2 immunity: review and applications to phase 3 vaccine candidates. Lancet 2020; published online October 13, 2020. https://doi.org/10.1016/S0140-6736(20)32137-1.

9. Liu H, Su D, Zhang J, et al. Improvement of pharmacokinetic profile of TRAIL via Trimer-Tag enhances its antitumor activity in vivo. Scientific Reports 2017;7, 8953.

10. Shi S, Zhu H, Xia X, et al. Vaccine adjuvants: Understanding the structure and mechanism of adjuvanticity. Vaccine 2019;37:3167–78.

11. Liang JG, Su D, Song T-Z, et al. S-Trimer, a COVID-19 subunit vaccine candidate, induces protective immunity in nonhuman primates. Preprint posted September 24, 2020 on BioRix https://doi.org/10.1101/2020.09.24.311027

12. Zogheri A, Di Mambro A, Mannelli M, Di Serio MG. Hyponatremia and pituitary adenoma: think twice about the etiopathogenesis. J Endocrinol Invest 2006;29:750–3.

13. Rajput R, Jain D, Pathak V. Recurrent hyponatremia as pPresenting manifestation of pituitary macroadenoma. 2017;44:46–49.

14. Bopeththa BVKM, Niyaz SMM, Medagedara C. Pituitary macroadenoma presenting as severe hyponatremia: a case report. J Med Case Rep 2019;13:40.Vaccine 2019;37:

15. Cohet C, Van der Most R, Bauchau V, et al. Safety of AS03-adjuvanted influenza vaccines: a review of the evidence. Vaccine 2019;37:3003–21.

16. Langley JM, Risi G, Caldwell M, et al. Dose-sparing H5N1 A/Indonesia/05/2005 prepandemic influenza vaccine in adults and elderly adults: a phase III, placebo-controlled, randomized study. J Infect Dis 2011;203:1729–38.

17. Mulligan MJ, Lyke KE, Kitchin N, et al. Phase I/II study of COVID-19 RNA vaccine BNT162b1 in adults. Nature 2020;586:589–93.

18. Jackson LA, Anderson EJ, Rouphael NG, et al. An mRNA vaccine against SARS-CoV-2 — preliminary report. New Engl J Med 2020;383:1920–31.

19. Barry M, Cooper C. Review of hepatitis B surface antigen-1018 ISS adjuvant-containing vaccine safety and efficacy. Exp Opin Biol Ther 2007;11:1731–7.

20. Halperin SA, Ward B, Cooper C, et al. Comparison of safety and immunogenicity of two doses of investigational hepatitis B virus surface antigen co-administered with an immunostimulatory phosphorothioate oligodeoxyribonucleotide and three doses of a licensed hepatitis B vaccine in healthy adults 18–55 years of age. Vaccine 2012;30:2556–63.

21. Moderbacher CR, Ramirez SI, Dan JM, et al. Antigen-specific adaptive immunity to SARS-CoV-2 in acute COVID-19 and associations with age and disease severity. Cell 2020;183: P996-1012.E19.

22. Okba NMA, Müller MA, Li W, et al. Severe acute respiratory syndrome coronavirus 2-specific antibody responses in coronavirus disease patients. Emerg Infect Dis 2020;26:1478–88.

23. Mulligan MJ, Lyke KE, Kitchin N, et al. Phase I/II study of COVID-19 RNA vaccine BNT162b1 in adults. Nature 2020;586:589–93.

24. Jackson LA, Anderson EJ, Rouphael NG, et al. An mRNA vaccine against SARS-CoV-2 – preliminary report. New Engl J Med 2020;383:1920–31.

25. Folegatti PM, Ewer KJ, Aley PK, et al. Safety and immunogenicity of the ChAdOx1 nCoV-19 vaccine against SARS-CoV-2: a preliminary report of a phase 1/2, single-blind, randomised controlled trial. Lancet 2020;396:467–78.

